# A mixed methods study of seasonal influenza vaccine hesitancy in adults with chronic respiratory conditions

**DOI:** 10.1101/2021.03.01.21252674

**Authors:** Lynn Williams, Karen Deakin, Allyson Gallant, Susan Rasmussen, David Young, Nicola Cogan

## Abstract

**Background:** Seasonal influenza vaccination is recommended for patients with chronic respiratory conditions, but uptake is suboptimal. We undertook a comprehensive mixed methods study in order to examine the barriers and enablers to influenza vaccination in patients with chronic respiratory conditions.

**Methods:** Mixed methods including a survey (n=429) which assessed socio-demographics and the psychological factors associated with vaccine uptake (i.e. confidence, complacency, constraints, calculation and collective responsibility) with binary logistic regression analysis. We also undertook focus groups and interviews (n=59) to further explore barriers and enablers to uptake using thematic analysis.

**Results:** The survey analysis identified that older participants were more likely to accept the vaccine, as were those with higher perceptions of collective responsibility around vaccination, lower levels of complacency, and lower levels of constraints. Thematic analysis showed that concerns over vaccine side effects, lack of tailored information and knowledge, and a lack of trust and rapport with healthcare professionals were key barriers. In contrast, the importance of feeling protected, acceptance of being part of an at-risk group, and feeling a reduced sense of vulnerability after vaccination were seen as key enablers.

**Conclusions:** Our findings showed that the decision to accept a vaccine against influenza is influenced by multiple sociodemographic and psychological factors. Future interventions should provide clear and transparent information about side effects and be tailored to patients with chronic respiratory conditions. Interactions between patients and their healthcare providers have a particularly important role to play in helping patients address their concerns and feel confident in vaccination.

## 1 BACKGROUND

The importance of vaccination and vaccine hesitancy have come into sharp focus due to the COVID-19 pandemic [1]. But the problem of vaccine hesitancy for other vaccines, including the annual seasonal influenza vaccine, is long established. The term ‘vaccine hesitancy’ refers to the ‘delay in acceptance or refusal of vaccines despite availability of vaccine services’ [2]. Patients with chronic respiratory conditions are particularly susceptible to serious complications that can arise from influenza, which can lead to hospitalisation [3]. The World Health Organisation (WHO) recommends annual seasonal influenza vaccination for people with long-term medical conditions, including those with chronic respiratory conditions, with a target uptake of 75% [4]. However, vaccination rates across Europe fall well below this [5]. The present study aims to understand the barriers and enablers to influenza vaccination in patients with chronic respiratory conditions.

The reasons for vaccine hesitancy are complex, involving individual influences, contextual influences and vaccine and vaccination-specific issues [6]. Increasingly, psychological factors are being recognised as providing the best explanations for people not taking up vaccinations [7]. Importantly, these factors are amenable to change through intervention [6]. To date, the research on influenza vaccine hesitancy in patients with chronic respiratory conditions has focussed largely on socio-demographic factors. For example, vaccination rates are higher in patients with comorbidities, while being male, younger in age, and a current smoker are all factors that have been associated with lower uptake [8]. However, the research linking influenza vaccine uptake to sociodemographic factors is inconsistent [7] and these factors cannot explain why there has been a rise in vaccine hesitancy over time. Evidence also suggests that psychosocial variables (e.g., risk perceptions) are better at explaining past vaccination behaviour than demographic, socioeconomic, and health variables [9].

In a systematic review on the barriers to influenza vaccination, psychological determinants including a lack of confidence (i.e. trust in the safety and effectiveness of the vaccine and in healthcare professionals and healthcare systems), inconvenience (the ease with which the individual can access the vaccine), and complacency (perceived risk of the illness and necessity of the vaccine), reflected in the ‘3C’ model of vaccine hesitancy [2], were related to uptake across risk groups [7]. More recently, two further ‘Cs’ have been suggested in an extension of the ‘3C’ model, with the ‘5C’ model also including calculation (i.e. engaging in information searching about the vaccine) and collective responsibility (vaccinating due to a sense of social responsibility) [10]. However, to-date, no study has undertaken a detailed examination of these psychological barriers to influenza uptake in patients with chronic respiratory conditions. Importantly, knowing the psychological barriers and enablers behind vaccination behaviour can inform the development of evidence-based vaccination policy and practice [11].

A further gap in the literature is that limited attempts have been made to integrate and synthesise qualitative findings on influenza vaccine hesitancy. Specifically in the context of chronic respiratory conditions, no studies have examined the reasons for vaccine hesitancy from a qualitative standpoint. The present study adopted a mixed methods approach, combining quantitative (survey) and qualitative methods (focus groups/ interviews). These methods provided complementary insights in order to develop a comprehensive understanding of the psychological barriers and drivers to influenza vaccine uptake in patients with chronic respiratory conditions.

## 2 METHODS

This exploratory sequential mixed methods study used quantitative (cross-sectional survey) and qualitative (focus groups and interviews) methods. Ethical approval was received from the institutional research ethics committee. Data collection was completed between May and October 2019, before the start of the COVID-19 pandemic.

### 2.1 Quantitative study

Overall, 429 participants with a chronic respiratory condition completed the survey (participant characteristics are shown in Table 1). We used convenience sampling with participants recruited to purposive criteria. In order to take part in the survey, participants had to be aged 18-64 and have a chronic respiratory condition, thus meeting the criteria to be offered an annual influenza vaccination free of charge by the National Health Service in the UK. Participants were recruited to the online survey via advertisements on social media and through partner organisations (including the Asthma UK and British Lung Foundation Partnership).

**Table 1.** Sociodemographic and health variables for the survey sample

| Variable |  | N | % |
| --- | --- | --- | --- |
| Gender | Female | 273 | 63.6 |
|  | Male | 149 | 34.7 |
|  | Missing | 7 |  |
| Education | High School | 115 | 27.1 |
|  | College | 134 | 31.5 |
|  | University | 109 | 25.6 |
|  | Postgraduate | 67 | 15.8 |
|  | Missing | 4 |  |
| Deprivation | SIMD 1 (most deprived) | 104 | 26.7 |
|  | SIMD 2 | 79 | 20.3 |
|  | SIMD 3 | 81 | 20.8 |
|  | SIMD 4 | 68 | 17.4 |
|  | SIMD 5 (least deprived) | 58 | 14.9 |
|  | Missing | 39 |  |
| Type of Respiratory Condition | Asthma | 346 | 80.7 |
|  | COPD | 82 | 19.1 |
|  | Other | 29 | 6.8 |
| Severity of Condition | Mild | 170 | 40.1 |
|  | Moderate | 208 | 49.1 |
|  | Severe | 46 | 10.8 |
|  | Missing | 5 |  |
| Influenza Vaccination Uptake | Yes | 240 | 55.9 |
|  | No | 189 | 44.1 |

#### 2.1.1 Key measures

Demographic and disease characteristics: Gender, age, type of respiratory condition, severity of condition, educational attainment, and socio-economic status (assessed by the Scottish Index of Multiple Deprivation (SIMD) [12].

Vaccination behaviour: Participants were asked if they had received the influenza vaccination in the previous 12 months (yes/no).

Barriers and enablers to vaccination: The 5C scale assesses psychological antecedents of vaccination and comprises five subscales measuring confidence (e.g., ‘I am completely confident that vaccines are safe’), complacency (‘vaccination is unnecessary because vaccine-preventable diseases are not common anymore’), constraints (‘everyday stress prevents me from getting vaccinated’), calculation (‘when I think about getting vaccinated, I weigh the benefits and risks to make the best decision possible’) and collective responsibility (‘I get vaccinated because I can also protect people with a weaker immune system’). Responses were measured on a seven-point scale (1= strongly disagree, 7= strongly agree) and scored by calculating the mean for each subscale [10].

#### 2.1.2 Statistical analysis

Univariate and multivariate binary logistic regression analyses were used to determine the sociodemographic (age, gender, educational attainment and deprivation), health (severity of condition) and psychological factors (confidence, complacency, convenience, calculation and collective responsibility) associated with vaccine uptake (0=not vaccinated, 1=vaccinated). All analyses were conducted using IBM SPSS Statistics (version 25) at 5% significant levels.

### 2.2 Qualitative study

A purposive sampling approach was utilised to recruit adults aged 18-64 years living in Scotland with a chronic respiratory condition. We sought to recruit those participants who over the previous five years had always vaccinated, occasionally vaccinated, and those who never vaccinated in order to understand the full range of vaccine hesitancy [2]. Participants for the qualitative component were recruited using the same methods as the survey participants, but were a distinct group. The focus groups and interviews were conducted by two researchers (KD and AG). In total, 59 participants took part in one of nine focus groups (n=38) or individual interviews (n=21). The mean focus group length was 49:27 minutes and mean interview length was 15:02 minutes. The mean age of participants was 42.8 (SD± 14.8) years and 70% of participants were female. Asthma was the most common type of chronic respiratory condition (72%). Of these participants, 24% had not vaccinated in the previous five years, 36% had occasionally vaccinated and 41% had always vaccinated.

Semi-structured focus groups and interviews were conducted using a topic guide developed for the purpose of this study: this covered experiences with the influenza vaccine, the factors influencing their decision whether or not to have it, and advantages and disadvantages of being vaccinated. Data from focus groups and interviews were audio-recorded and transcribed verbatim. Transcripts were anonymised and pseudonyms were created for each participant. Transcripts were imported and managed in NVivo 12. The data were analysed in accordance with inductive thematic analysis in order to address the exploratory aims of the qualitative component of the study [13]. Preliminary themes created by the researcher (KD) were cross-checked by a co-researcher (NC) to offer a further credibility check of the emergent themes. Quotes from the transcripts that captured discrete aspects of each theme were identified.

### 2.3 Integration of findings

To integrate the findings of the quantitative and qualitative components, we first entered the key results of each component into a table (see Table 4). Drawing on the constant comparative method [14] areas of similarity and difference in the findings were then highlighted by one of the researchers (LW), who generated interpretative statements for the overall meaning of the two contributing studies. These statements were then checked by the rest of the team to ensure their validity and coherence.

**Table 4.** Integration of the main findings across the quantitative and qualitative studies

| Quantitative study | Qualitative study | Integration and synthesis |
| --- | --- | --- |
| Older participants were more likely to get vaccinated |  | The quantitative findings suggest particular targeted efforts may be needed for younger adults with chronic respiratory conditions |
| Having greater confidence in vaccines was associated with getting vaccinated (univariate analysis) | The perception that the vaccine causes adverse side effects as well as a lack of trust and rapport in general with HCPs was identified as a barrier | The importance of having trust in vaccines and the system that delivers them was identified in both components. Addressing concerns about side effects through transparent communication with healthcare providers may be important |
| Higher levels of complacency about vaccination were associated with not getting vaccinated | Acceptance of being at greater risk due to respiratory condition and the belief that influenza represented a significant threat to health' leading to feelings of being protected, was a facilitator to vaccination | Both components identified the key role for an individual's perception of risk. Those who perceived influenza to be a significant threat and that they were at greater risk for it were more likely to get vaccinated. |
| Higher perceptions of constraints around vaccination were associated with not getting vaccinated | Lack of knowledge about the vaccine was identified as a barrier, with participants wanting more detailed information tailored to their condition | Practical constraints were identified as being important in both components. The provision of more tailored information may be useful. |
| Higher levels of collective responsibility were associated with getting vaccinated | Participants mentioned the importance of protecting vulnerable people that they were in contact with | The willingness to protect others was identified in both components. Emphasising that getting vaccinated can protect others, including vulnerable friends and family, may be useful. |

## 3 RESULTS

### 3.1 Quantitative study

#### 3.1.1 Sample characteristics

The sample comprised 429 participants (63.6% female) with a mean age of 41.8 years (SD = 13.7).

Most reported post-secondary school education (72.9%). In terms of deprivation, 26.7% lived in the most deprived areas, based on SIMD quintile. The majority of participants reported having asthma (80.7%), 19.1% reported having COPD, and 6.8% reported having another respiratory condition. Regarding severity, 39.6% reported that their condition was mild, 48.5% reported that it was moderate, and 10.7% that it was severe.

#### 3.1.2 Factors associated with influenza vaccination uptake

Binary logistic regression analysis compared those who had received the influenza vaccine in the previous 12 months (n=240; 55.9%) with those who had not (n=189; 44.1%). Univariate analyses showed that there was a significant effect of age, severity of condition, confidence, complacency, constraints and collective responsibility on uptake of the influenza vaccination. There was no effect of gender, educational attainment, deprivation, or calculation (see Table 2).

**Table 2.**
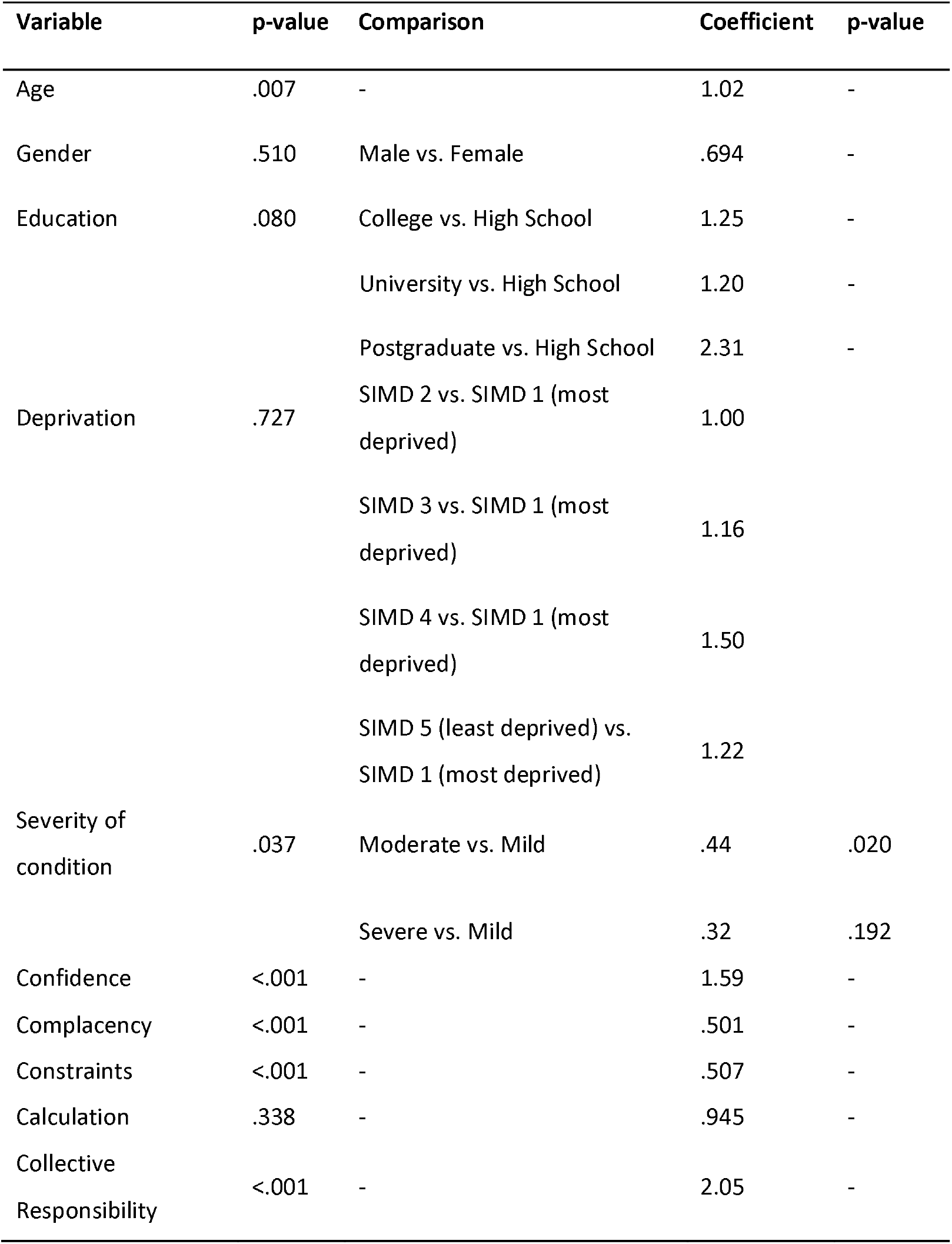
Univariate analysis of influenza vaccination uptake

For the multivariate logistic regression, we entered those variables that were significant in the univariate analysis (i.e. age, severity of condition, confidence, complacency, constraints, collective responsibility). Age, complacency, constraints, and collective responsibility remained significantly associated with uptake in the multivariate analysis, but severity of condition and confidence were no longer significant. In the final model, vaccination uptake was associated with being older and having higher levels of collective responsibility, lower levels of complacency, and lower perceptions of constraints to vaccination (see Table 3).

**Table 3.** Multivariate analysis of influenza vaccination uptake

| Variable | p-value | Comparison | Coefficient |
| --- | --- | --- | --- |
| Age | .030 | - | 1.02 |
| Severity of condition | .118 | Moderate vs. Mild | 1.395 |
|  |  | Severe vs. Mild | 2.42 |
| Confidence | .060 | - | 1.19 |
| Complacency | .008 | - | .72 |
| Constraints | <.001 | - | .62 |
| Collective Responsibility | .001 | - | 1.52 |

### 3.2 Qualitative study

The inductive thematic analysis highlighted seven themes, four reflecting barriers to vaccination uptake and three reflecting enablers to uptake. The themes that reflected barriers to vaccination uptake were evident among all participants who were occasional or non-vaccinators. The themes that reflected enablers to vaccination were evident among all those who were regular vaccinators.

#### 3.2.1 Barriers to influenza vaccination uptake

##### Lived experience of perceived adverse side-effects

Participants who did not routinely vaccinate reported that they believed the vaccine caused adverse side-effects. For some, this belief was based on personal experience. Matthew (Non-Vaccinator) described it as ‘*the worst I’ve ever felt in my life… I was ill for about ten days. I couldn’t get out of my bed for ten days. I was really bad’*. Belinda added:

> *I usually get really fatigued, and it’s, I get like flu-like symptoms, in response to the jag. And that will go on for a few weeks. So, the past sort of two or three years, I’ve not bothered to get it, just to avoid feeling unwell after it (Belinda, Occasional Vaccinator).*

Participants also described the possibility of adverse effects of the vaccine being too great a risk:

> *I had very mixed thoughts on the flu jag. I understand the background of it. I understand it’s to stop me getting ill and actually having a disease that would kill me but I still felt very vulnerable taking it that it would in fact push me… make me feel unwell (Erin, Non-Vaccinator).*

Many participants rejected the vaccine because they witnessed others experience perceived adverse effects. For example, Heather was certain that she did not want the vaccination because of her mother’s experiences:

> *I’ve never had the flu jag and I don’t intend getting the flu jag. Everybody I know that gets it, gets the flu and gets it badly. My mother used to get it and she was ill for weeks after it (Heather, Non-Vaccinator).*

##### Uncertainty and mistrust in vaccination

Participants described their lack of knowledge about the vaccine’s ingredients and how they work. Participants reported that this uncertainty hindered their trust in the vaccination. Victoria (Non-Vaccinator) stated: *‘… if they told you more information it might actually help you take some trust in it*.’ Another participant added:

> *Uncertainty, I don’t know, like, thinking about what it is that I’m putting into my body and thinking about, like, do I need this, do I not need this, trying to weigh up those things, and I don’t really know, I suppose, yeah, you don’t really know (Kathleen, Non-Vaccinator).*

Participants voiced their frustration at not being given *detailed* information as to *how and why the vaccination worked*; consequently they felt unable to make an informed decision about whether to vaccinate:

> *I think just that sort of one-size-fits-all thing that they kind of throw on you, I think that really needs to be kind of, I guess, adjusted to each person’s needs (Stacey, Non-Vaccinator).*

Participants believed that they had not been provided with enough information about the vaccine and, for some, this appeared to foster a feeling of mistrust:

> *So, why not give us the correct information or why not make that little leaflet, that they’re obviously sending out to us in mass production, just have a little bit more information in it? Give a little bit more about what the jab actually is, what’s the success rate? (Victoria, Non-Vaccinator).*

##### Questioning the need for the vaccine

Many participants who chose not to vaccinate did not believe it would protect them from influenza:

> *I don’t feel that the year I got it [the vaccine] and every year subsequently it has had any impact on that… So I seem a bit unconvinced by its ability to work (Alastair, Non-Vaccinator).*

Some participants did not believe that influenza represented a serious threat to their health. For example, Belinda stated:

> *I think, like for me, it’s just, I feel like, if I got the flu, my body would be able to handle it (Belinda, Occasional Vaccinator).*

Participants often considered the severity of their respiratory condition when assessing their vulnerability to influenza. Ross stated:

> *…As I say personally I just, you know, I don’t suffer like other people do with getting the flu. Probably got mild COPD, you know, I don’t use inhalers or anything like that. I’m offered it consistently, but until I get a bad flu and maybe get very ill as a result, I would probably take it then obviously, but just don’t feel the need at the moment (Ross, Non Vaccinator).*

##### Lack of trust and rapport with healthcare professionals (HCPs)

Participants who did not vaccinate, or had only occasionally done so, felt that their relationship with HCPs lacked several qualities they valued, for example, not recognising them as an individual or listening to their concerns. This was expressed by Dianne in the following way:

> *I was with the same doctor for, well, since I’ve been born, basically, but they changed practice about three years ago. And I used to get the same nurse, and she knew everything about you. But now, I’d say, I don’t even know their names. Or the doctor’s name, actually, you know. (Dianne, Occasional Vaccinator).*

Participants also reported that communication about the vaccination had not been tailored to their needs or level of understanding:

> *I feel like it’s just been a leaflet and that’s kind of it. I don’t think I’ve ever asked, you know, what is it that goes into this, tell me the actual ingredients in a layman’s way that I’m going to understand. I feel like if I did ask, I’m not sure how well they’d actually be able to tell me as well (Stacey, Non-Vaccinator).*

A few participants perceived their relationship with HCPs to be particularly strained. Stacey described feeling pressured to accept the vaccination whilst attending an appointment for her annual asthma check-up:

> *[It’s] stressful as well when you’re kind of up against a doctor and asthma nurses telling you to kind of constantly go [to get the vaccine] and you’re kind of having to stand your corner every time (Stacey, Non-Vaccinator).*

Another participant, Rosie, described her shock when her GP had prepared the vaccination in advance of her appointment, without asking her if she would like it:

> *One time, I went for an appointment, to see the doctor about something entirely different, and the doctor was sitting there with a needle, all prepared, ready for me to walk in, because it was that time of year. So, I feel as though we’ve not really got a lot of choice, unless you do stand up for yourself against the doctor. Because that doctor, that day, was willing to just go ahead, whether I said yes or no, until I said, well, stop, and that was it (Rosie, Non-Vaccinator).*

Perceived undue pressure from HCPs exacerbated participants’ feelings of uncertainty about the vaccine and of being undermined as autonomous individuals.

#### 3.2.2 Enablers to influenza vaccination uptake

##### Feeling cared for and valued

There was a clear sense from participants who vaccinated that they felt their health needs were being catered for. They described the influenza vaccination as easy to get because their GP practice provided a variety of ways for them to obtain it:

> *So we get a letter sent to us from our surgery telling us the date that they’re doing the mass vaccination and all the doctors and nurses attend the surgery, [they] give up their time… [it] sticks in my mind … because it’s like our surgery, it’s like a community thing, is you actually see doctors and nurses with smiles on their faces because they’re officially not at work or anything like that. They’re just there doing something and they were happy… (David, Regular Vaccinator).*

Several participants explained that their vaccination was administered by a nurse with whom they were familiar and who showed their concern for them by prompting them to be vaccinated during routine visits. Feelings of being cared for and valued engendered participants’ trust in the advice of their HCP:

> *… the nurse is always really nice and jolly and you usually get a good laugh when you go, bit of banter with the nurse, and they always make you feel so comfortable, so I’ve not really thought. I don’t feel nervous or anything about it, it’s just part of … it is what it is and you just do it and that’s it (Nina, Regular Vaccinator).*

Participants reported having been provided with a variety of vaccination opportunities, including at routine check-ups, mass vaccination events or weekend appointments; this resulted in participants gaining a sense of feeling cared for, valued and that their wellbeing mattered:

> *I think they probably make it easier for me to get it, and that’s what encourages me to get it. I think if nobody was phoning me, reminding me that I needed to go and get it … the fact that somebody is reminding me to go and get it, is definitely a huge influencer, the fact that I get it. I think if it was just left up to me to do it on my own will, I don’t know that it would happen (Kathleen, Occasional Vaccinator).*

##### Acceptance of advice to vaccinate given ‘at risk’ status

Participants who vaccinated had been told by their HCPs that they were at greater risk of contracting influenza and they were comfortable accepting this advice to vaccinate. Lawrence (Regular Vaccinator) described the vaccination as a *‘necessity*’ and ‘*required*’. He added, ‘*I’ve got this illness so, I’m only taking professional advice*’. Similarly, another participant said:

> *They wouldn’t offer me it if they didn’t think I needed it so that’s pretty much my feelings about it but it’s obviously something that they think I should have so I’m quite happy to go with that … to follow their advice (Michelle, Occasional Vaccinator).*

Having trust in the expertise of their doctor and the scientists who developed the vaccination facilitated their decision to vaccinate.

> *I don’t really know about the vaccine itself, or about the immunity that it gives you. To be honest, I’m quite ignorant about what they’re actually doing putting it into you; I’m just at the hands of the GP or whoever it is that gives you it … I trust in them (Alexa, Regular Vaccinator).*

##### Reduced sense of vulnerability and the importance of protection

Participants who chose to uptake the vaccination described how it helped reduce their sense of physical vulnerability; influenza represented a significant threat to their health:

> *I’m very compromised so if I was to get the flu… my lungs wouldn’t cope with it. I think I just wouldn’t … my body wouldn’t be able to cope with it (Erin, Occasional Vaccinator).*

Participants described the vaccination as giving them ‘*peace of mind*’, a sense of being ‘*safer*’ (Oliver, Regular Vaccinator) and ‘*relief*’ at being protected against influenza (Jill and Anne, Regular Vaccinators). Nina (Regular Vaccinator) described the vaccination as instilling her with ‘*confidence*’ in her health. Similarly, another participant said:

> *As long as it continues to help me to live, which is more like, thank God I’m not going to be stuck in my bed for weeks and all the rest of it, I will continue to take my flu jag. I just think thank God there is such a thing as the flu vaccinations (Jane, Regular Vaccinator).*

Participants also noted that the sense of protection extended to those around them as well:

> *Yeah, definitely, that sort of help the immunity thing. I work for a children’s palliative care provider, so who we work for is, obviously, it’s terminal children, and very complex conditions. So that sort of immunity, from that point of view, I think is really important… (Scott, Regular Vaccinator).*

## 4 DISCUSSION

The current study is the first to examine the demographic and psychological barriers and enablers to influenza vaccination in adults with chronic respiratory conditions. Through our mixed methods approach we examined the psychosocial patterning of vaccine uptake and the psychological processes that influence the decision to vaccinate. The integration of the key findings from the quantitative and qualitative components is shown in Table 4.

These findings demonstrate that having trust in vaccines and HCPs is key. In particular, addressing patients’ concerns about side effects through transparent communication may be useful in building trust and confidence [15]. The role of complacency was also important: those individuals who perceived influenza to be a serious illness, and who accepted they were at greater risk, were more likely to vaccinate. Participants also noted feeling protected and less vulnerable following vaccination. Practical constraints were also important, with participants emphasising the importance of the information that is made available to them. Many wanted more detailed information that was tailored to their respiratory condition [16]. The survey findings also suggest that having a greater sense of collective responsibility is important, and intervention efforts could aim to foster this by explaining that being vaccinated can protect others. Our findings also indicate that it may be important to target intervention efforts towards younger adults with respiratory illnesses, as they are less likely to get vaccinated.

In terms of intervention development, our findings suggest there are multiple factors that can be targeted in order to improve vaccine uptake. The key role that HCPs can play in shaping their patients’ vaccine decision making was highlighted [17]. Therefore, the development of training and time-efficient strategies for HCPs around how to communicate with vaccine-hesitant patients is warranted [18, 19].

Strengths of the study include the mixed methods approach which has allowed us to build a comprehensive picture of the barriers and enablers to influenza vaccine uptake. In addition, our recruitment approach ensured that we recruited participants from across the vaccine hesitancy continuum, including those who were non-vaccinating, those who vaccinated occasionally and those who vaccinated regularly. Limitations include that our findings are limited to participants living in Scotland with a chronic respiratory condition, and so may not be generalisable to other chronic health conditions or national contexts. In addition, as our data was collected prior to the COVID-19 pandemic we do not know what impact the pandemic may have on vaccination beliefs and behaviour in general.

## Conclusion

Our mixed methods study identified a number of sociodemographic and psychological factors that influence influenza vaccination uptake in adults with chronic respiratory conditions. Addressing issues of complacency around the vaccination, building confidence and trust in the vaccine through transparent communication about side effects, overcoming practical constraints including the provision of tailored information, and emphasising the importance of collective responsibility can all be utilised in order to increase vaccine uptake. In addition, targeted interventions for younger adults with chronic respiratory conditions are needed.

## Data Availability

The data are not publicly available due to ethical or privacy restrictions

## Acknowledgements

We would like to thank the participants that took part in the study and the Asthma UK and British Lung Foundation Partnership for their support with participant recruitment.

